# Non-specific effects of vaccines on all-cause mortality: a meta-analysis of randomized controlled trials (RCTs) 2012-2025

**DOI:** 10.64898/2025.12.31.25343212

**Authors:** Josephine Schou, Anders Hviid, Eskild Petersen, Lone Simonsen

## Abstract

**Background:** Early observational studies reported substantial reductions in all-cause mortality, suggesting that childhood vaccines might influence health in ways beyond their targeted disease. Such Non-Specific Effects (NSEs) of vaccines could potentially have important implications for the Expanded Programme of Immunization (EPI). In 2014 World Health Organization (WHO) Strategic Advisory Group of Experts (SAGE) commissioned a review of the NSE evidence, which called for more Randomized Controlled Trials (RCTs). We aimed to review all RCTs published after this WHO-commissioned review.

**Methods:** Following PRISMA guidelines, we searched systematically for RCTs published during 2012-2025. Inclusion criteria: children under 5 years and all-cause mortality as the primary outcome for any vaccine intervention. Risk estimates were pooled using a random effects model, reflecting the underlying hypothesis: that live-attenuated vaccines greatly reduce, and inactivated vaccines increase all-cause mortality.

**Results:** We identified seven RCTs that fit the inclusion criteria, all of them of live-attenuated vaccines (Bacillus Calmette-Guérin, measles, and oral polio vaccine). In total, 52,596 participants were included across these studies. Five were conducted in Guinea-Bissau, one in Guinea-Bissau and Burkina Faso, and one in India. The RCTs were not blinded, and no placebo was used, consistent with a moderate risk of bias. Our meta-analysis found no significant evidence of NSEs (vaccine effectiveness 5%; −16% to 22%). One study of low birthweight newborns admitted to neonatal intensive care units in India found a small but statistically significant NSE (17%; 2% to 31%); interestingly the effect primarily played out in the first 3 days of the study, raising concern of bias.

**Discussion:** The meta-analysis of seven RCTs of live-attenuated vaccines found no evidence of significant NSE on all-cause mortality. This was also true when adding RCTs from before 2012 studying NSEs. In conjunction with the WHO-SAGE review, the current body of evidence on NSEs on all-cause mortality does not warrant a paradigm shift, nor does it provide a rationale for changing vaccine programs.

## Introduction

Non-specific effects (NSEs) of vaccines - meaning beneficial or detrimental effects on health outcomes beyond protection against the targeted disease - have been the focus of debate and investigation for several decades. The underlying hypothesis is that live attenuated vaccines (such as Bacillus Calmette-Guérin (BCG), measles vaccine (MV), oral polio vaccine (OPV)) have beneficial NSEs, while inactivated vaccines (such as diphtheria-tetanus-pertussis (DTP)) have negative NSEs (see for example (1)).

Early observational studies reported striking NSEs; for example, one study found a 37% reduction in all-cause mortality among children who received the measles vaccine early rather than later in infancy (2). Because this is an enormous claim, and because observational studies are inherently prone to bias (3), there has been a drive towards studying such effects in randomized controlled trials (RCTs) to enable an unbiased assessment of any effect between vaccines and all-cause mortality, and to assess potential causality.

Although RCTs are in principle unbiased, there are ethical constraints to using blinding and placebo controls in these study designs, because the vaccines are already adapted for use. Thus, all NSE trials conducted after the implementation of the Extended Programme on Immunization (EPI) in 1974 are not blinded and have no placebo group, introducing a moderate risk of bias. Given the potentially far-reaching implications of NSEs, substantial efforts have been taken to evaluate relevant studies. The WHO Strategic Advisory Group of Experts (SAGE) commissioned a comprehensive review, published in 2014, of all observational and randomized studies, encompassing evidence available up to November 2012 (4). Overall, the WHO review identified a high risk of bias across study designs, including both RCTs and observational studies. While RCTs investigating MV and BCG were available, no results from RCTs of DTP had been published at the time of the review. Although the WHO-commissioned assessment indicated that MV and BCG might reduce mortality not related to measles and tuberculosis, the review expressed low confidence in these estimates due to study limitations and pervasive risk of bias. Consequently, the review concluded that the existing evidence was insufficient to warrant changes to immunization practices, and emphasized the need for additional RCTs, particularly concerning DTP (5).

Since the WHO-commissioned review, NSEs have been proposed to be a paradigm-shift in vaccinology (1). Proponents claim that NSEs could account for as much as halving all-cause mortality, which would be far greater than the direct effect of the vaccine on the targeted disease. This phenomenon, they say, should therefore impact the sequence of vaccinations (1). In this review, we aimed to provide an updated assessment of the body of evidence on NSEs by reviewing all RCTs published since the WHO-commissioned review in 2014. We included RCTs on any childhood vaccine published during 2012 to 2025 that assessed all-cause mortality as a primary endpoint for any vaccine intervention during the first 5 years of life. Building on the NSE hypotheses that proposes opposite mortality effects of live-attenuated and killed vaccines, we planned to pool results from the trials accordingly.

## Methods

We conducted a **review** of non-specific effects (NSEs) on all-cause mortality, focusing on studies published in English-language peer reviewed journals published after the WHO review by Higgins et al in 2014 (4). Our search was aimed at identifying RCTs published between January 2012 and December 2025 that met our predefined inclusion criteria. The search strategy was refined iteratively during pilot testing to account for alternative terminology (e.g., “heterologous”, “unspecific”) and ensure comprehensive coverage of relevant studies. The review adhered to the **PRISMA guidelines** for systematic reporting (6). We registered our protocol for this review with PROSPERO (CRD420251129006).

### Sources and Search Strategy

Searches were conducted across **PubMed, SCOPUS, Web of Science, EBSCOhost**, and **Cochrane** databases, using consistent Boolean operators and phrase-search syntax. To ensure an exhaustive search, we employed truncation and iterative refinement techniques. We also performed **Chain searches** to identify additional relevant review studies; the reference lists in such reviews were then perused to discover any additional RCTs that would fit our inclusion criteria. We also screened **Clinical trial IDs** to prevent duplicate reporting and ensure consistency between protocols and the reporting of primary findings. Searches were conducted in September and October 2025, with a final search on December 9, 2025, to ensure completeness and replicability. We conducted the first two searches manually, while the last search was done using the review platform, **Covidence** (www.covidence.org), as recommended by an information specialist. A detailed record of both manual searches can be found in Supplementary S1, tables S1.1 and S1.2; subsequent reporting in the article is based on the final search conducted in Covidence. Both manual and Covidence-assisted searches yielded the same 7 studies. Meta-analyses found through the searches were assessed for citations of primary RCTs, and the meta-analyses were subsequently excluded in this review. All searches and screenings were mainly conducted by JS with support from the other authors.

### Inclusion and Exclusion Criteria

Eligible studies were RCTs published between January 1^st^, 2012, and December 9^th^, 2025, that assessed NSEs of vaccines in children under 5 years of age, with all-cause mortality as the primary endpoint. Any vaccine-intervention was an inclusion criterion, as well as any randomization method – either individual, block or cluster. Studies were excluded if they were non-RCTs, discontinued RCTs, animal studies, natural experiments, or re-analyses/pooled studies, to avoid second-order synthesis. Trial registration details were cross-checked with reported outcomes and study periods to confirm congruency.

### Identification of studies

The latest search as of December 2025 yielded 240 hits. Of these, 158 duplicates were identified: 10 manually and 148 through Covidence. We screened 82 studies based on title and abstract, out of which we excluded 61 of them. We read 21 articles in full text. Seven of these were excluded as they were meta-analyses and reviews and only used for further citation searches. Of the 14 remaining articles, we excluded 7 studies, leaving a total of seven articles included in this review; see result section for the rationale for these exclusions. For the remaining 7 RCTs we used clinical trial IDs to verify the protocols and the registered primary outcomes using information from clinicaltrials.gov (6 studies) and Clinical Trial Registry – India (1 study).

### Pooled Data analysis

We treated all effect estimates the same, irrespective of whether they reported a hazard ratio (HR) or mortality risk ratio (MRR) as a measure of relative risk, regardless of follow-up duration. These estimates are presented as an Effect Size (ES) corresponding to relative risk (HR or MRR), following the approach used in the WHO review (4). ES data from each study were extracted by JS, and cross-checked by two authors, JS and LS, in a plenary session. We calculated the vaccine effectiveness (VE) as 1-ES.

The analysis was conducted independently by two of us, AH and JS, reaching the same result. We used a random-effects model as we anticipated heterogeneity across studies due to differences in population, setting, and vaccine interventions. Confidence intervals (95%CI) for the pooled effect size estimates were calculated using the Hartung-Knapp-Sidik-Jonkman (HKSJ) method, which provides more reliable and conservative control of type I error rates, especially when the number of studies is small (see Supplement S2 for *R meta* code).

## Results

We searched Scopus, PubMed, EBSCOhost, Web of Science and Cochrane Library up to December 9^th^, 2025, and performed a citation search based on seven review articles. From this we identified a total of 240 articles, then removed 158 that were duplicates. The remaining 82 papers were screened for title and abstract, on which basis we excluded 61 of them. The remaining 21 articles were screened in full text, leading to the exclusion of an additional 14 articles, leaving a total of 7 studies included in this review. The 14 articles were excluded for the following reasons: two because the study design was not RCT (7,8), one because it has already been included in the WHO-commissioned review (9), one because it was in fact a review (labeled “wrong publication type” in Figure 1) (10), and three articles because they did not have all-cause mortality as a primary outcome (11–13). Finally, 7 were excluded as they were meta-analyses or reviews (14–20).

**Figure 1.**
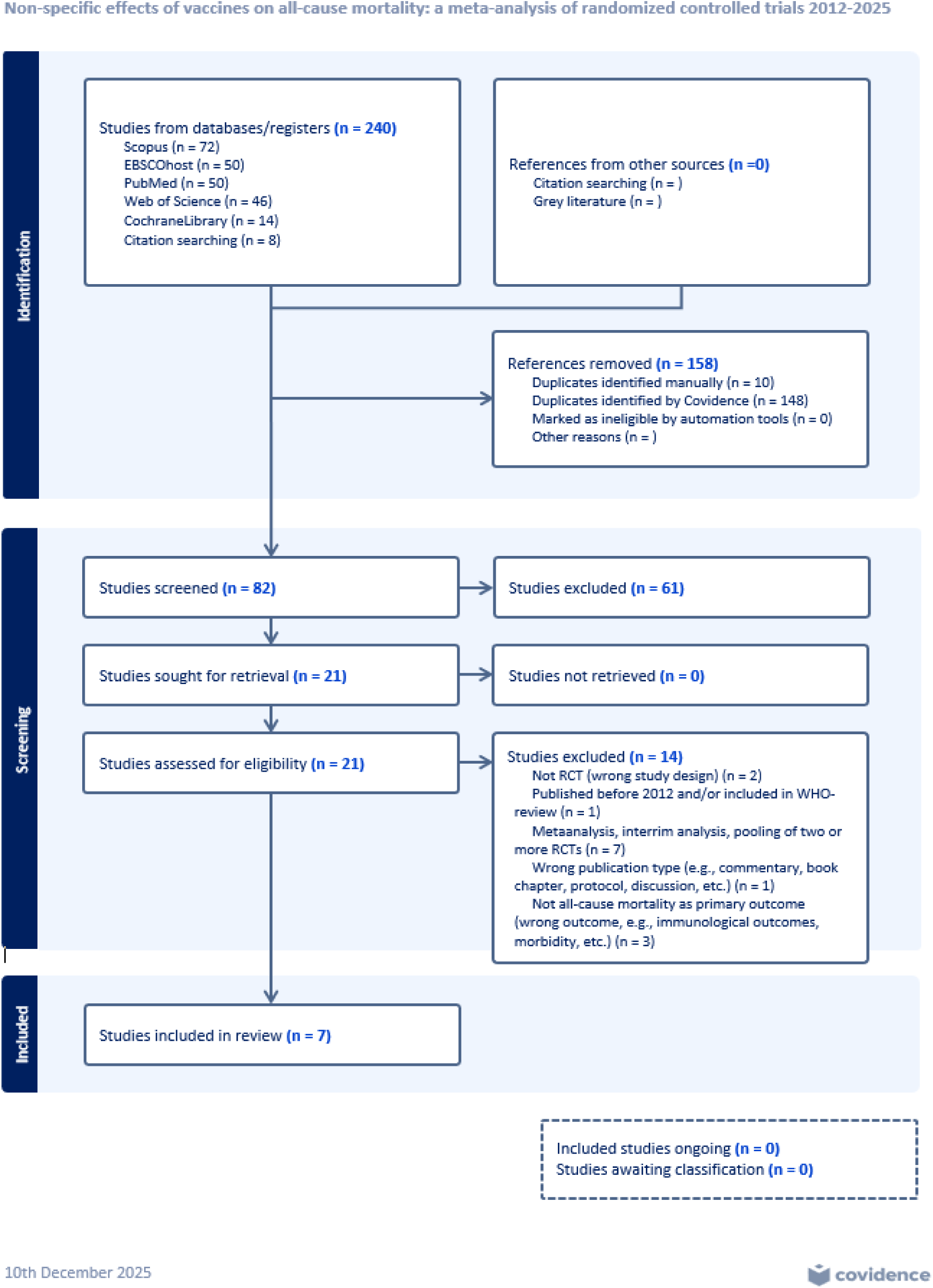
PRISMA flowchart of the inclusion process. Provided through Covidence.

### Included studies

Seven studies met the inclusion criteria, comprising a total of 52,596 participants (Table 1). Of these seven studies, six were conducted in West-Africa: five in Guinea-Bissau and one multicenter trial across Guinea-Bissau and Burkina Faso, while one study was set in Southeast India. All seven studies enrolled children of varying age ranges, ranging from neonates (of low or normal birth weight) to nearly 5-year-old children. Two trials enrolled low birth weight (LBW) neonates (21,22), one trial enrolled neonates (23), and one trial included children aged 4-7 months (24). Another trial enrolled children ages 4-7.5 months (25), while two trials enrolled children aged 9-35 months (26) and 9-59 months (27), respectively.

**Table 1.**
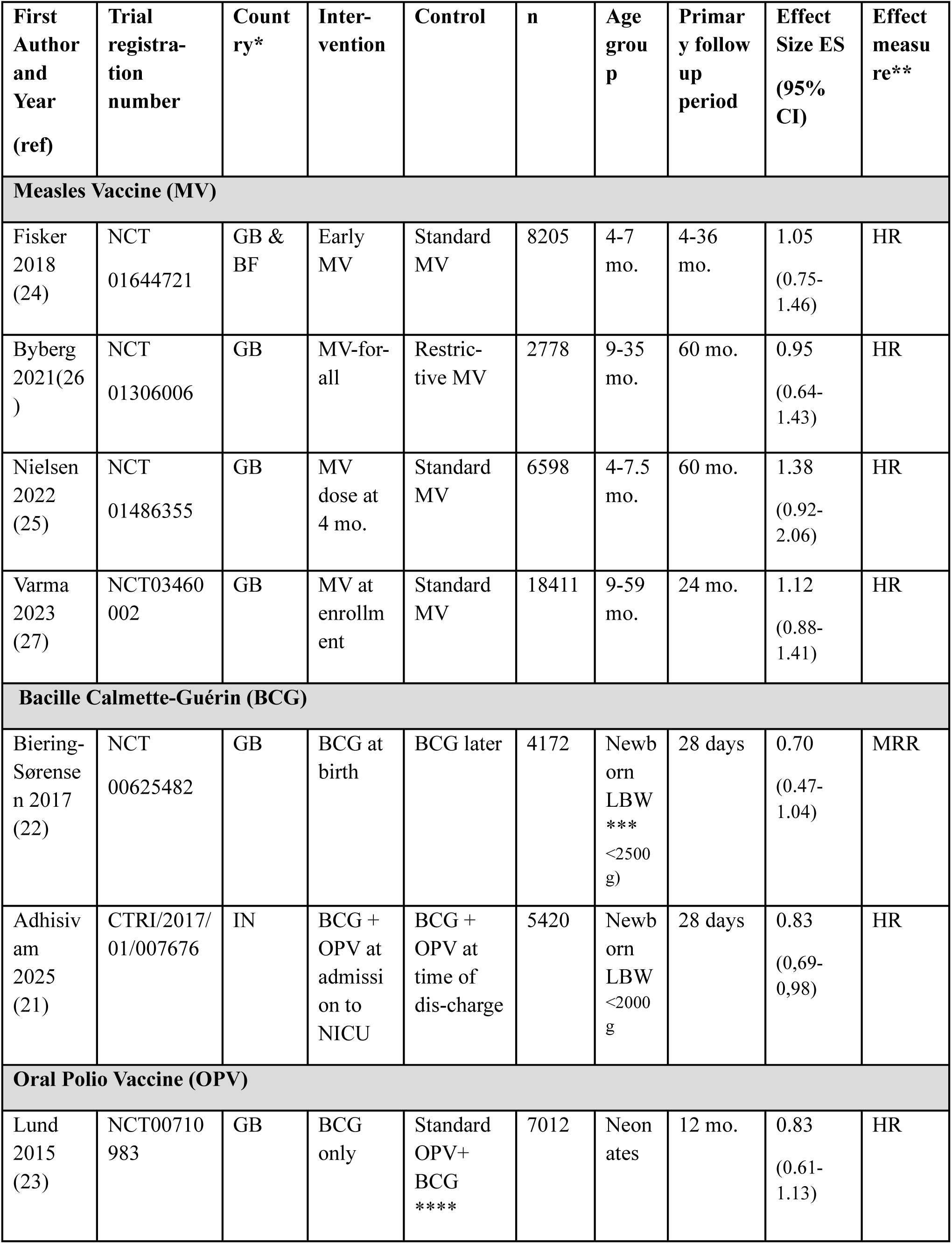
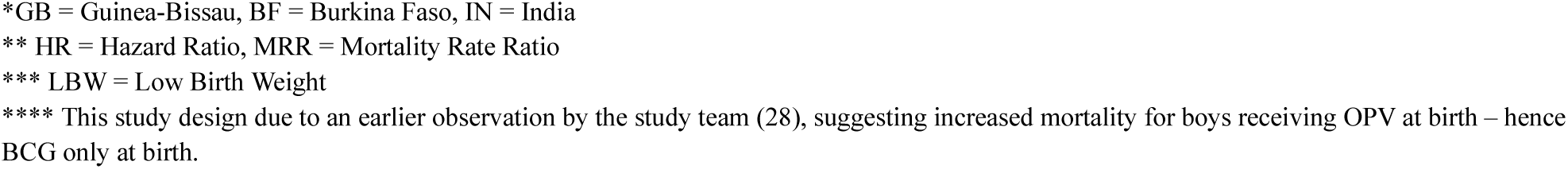
A list of the 7 included Randomized Controlled Trials (RCTs) on non-specific vaccine effects (NSE) using all-cause mortality as the primary outcome and published during 2012-2025. Two studies were cluster-randomized (26,27) and one study was block-randomized (22).

| First Author and Year (ref) | Trial registration number | Country* | Intervention | Control | n | Age group | Primary follow up period | Effect Size ES (95% CI) | Effect measure** |
| --- | --- | --- | --- | --- | --- | --- | --- | --- | --- |
| **Measles Vaccine (MV)** |  |  |  |  |  |  |  |  |  |
| Fisker 2018 (24) | NCT01644721 | GB & BF | Early MV | Standard MV | 8205 | 4-7 mo. | 4-36 mo. | 1.05 (0.75-1.46) | HR |
| Byberg 2021(26) | NCT01306006 | GB | MV-for-all | Restrictive MV | 2778 | 9-35 mo. | 60 mo. | 0.95 (0.64-1.43) | HR |
| Nielsen 2022 (25) | NCT01486355 | GB | MV dose at 4 mo. | Standard MV | 6598 | 4-7.5 mo. | 60 mo. | 1.38 (0.92-2.06) | HR |
| Varma 2023 (27) | NCT03460002 | GB | MV at enrollment | Standard MV | 18411 | 9-59 mo. | 24 mo. | 1.12 (0.88-1.41) | HR |
| **Bacille Calmette-Guérin (BCG)** |  |  |  |  |  |  |  |  |  |
| Biering-Sørensen 2017 (22) | NCT00625482 | GB | BCG at birth | BCG later | 4172 | Newborn LBW *** <2500 g) | 28 days | 0.70 (0.47-1.04) | MRR |
| Adhisivam 2025 (21) | CTRI/2017/01/007676 | IN | BCG + OPV at admission to NICU | BCG + OPV at time of discharge | 5420 | Newborn LBW <2000 g | 28 days | 0.83 (0.69-0.98) | HR |
| **Oral Polio Vaccine (OPV)** |  |  |  |  |  |  |  |  |  |
| Lund 2015 (23) | NCT00710983 | GB | BCG only | Standard OPV+ BCG **** | 7012 | Neonates | 12 mo. | 0.83 (0.61-1.13) | HR |

\*GB = Guinea-Bissau, BF = Burkina Faso, IN = India
\*\* HR = Hazard Ratio, MRR = Mortality Rate Ratio
\*\*\* LBW = Low Birth Weight
\*\*\*\* This study design due to an earlier observation by the study team (28), suggesting increased mortality for boys receiving OPV at birth – hence BCG only at birth.

Similarly, there was considerable variation in the follow-up periods which ranged from 28 days to the child’s 5^th^ birthday (60 months). We leaned on the stated primary follow-up time, even though in some studies the follow-up period was extended and several endpoints reported as a result.

All trials tested the hypothesis of NSEs of childhood vaccines on all-cause mortality. However, they investigate the hypothesis from somewhat different angles. For example Lund et al. (23) is sex-stratified based on earlier observations of OPV, suggesting increased mortality for boys receiving OPV at birth (13). Another trial, Byberg et al. (26), studied a measles-vaccine intervention with standard care is the control. The trial assessed whether a restrictive measles vaccination policy, implemented to minimize vaccine wastage, leads to higher child mortality by creating missed vaccination opportunities, compared with a policy offering measles vaccination to all unvaccinated children irrespective of age or group size.

As our review was designed to assess any vaccine intervention, we included the 7 RCTs presented in Table 1 in our meta-analysis, as they were all testing live-attenuated vaccines (BCG, OPV, and MV) and with a similar hypothesis expecting a huge reduction in all-cause mortality. We did this despite the study designs varying greatly (see above and table 1). The intervention was typically administered early or in accordance with the EPI schedule - at birth (BCG, OPV), at 6-14 weeks (OPV), and at 9 months (MV). The control groups received standard care. Six of the seven RCTs reported a Hazard Ratio (HR) as the Effect Measure, while one reported Mortality Rate Ratio (MRR). Four of the studies were randomized at the individual level (21,23–25), while two were cluster randomized (26,27) and one was block randomized (22).

We used the RoB-2-tool to assess the risk of bias within each RCT study, following the guidelines to the assessment tool (29). JS and LS assessed the seven included articles, without any disputes emerging. All seven study designs raised *some level of concern* of bias, primarily due to their open-label nature (unblinded randomization) and the absence of a placebo group. The results of these bias assessments for each study can be found in Supplement File (S3, RoB-2 assessments). Additional concerns were noted due to the concurrent administration of other vaccines or vaccine campaigns that occurred during trials, representing deviations from the intended interventions (RoB-2, Domain 2).

### Meta-analysis of Effect sizes in the seven RCTs

Only one of the seven RCTs demonstrated a significant reduction in all-cause mortality in the intervention group (ES<1), while the remaining six studies showed 95% CIs that overlapped with ES=1 (indicating no significant difference between the intervention and control groups) (Figure 2).

**Figure 2.**
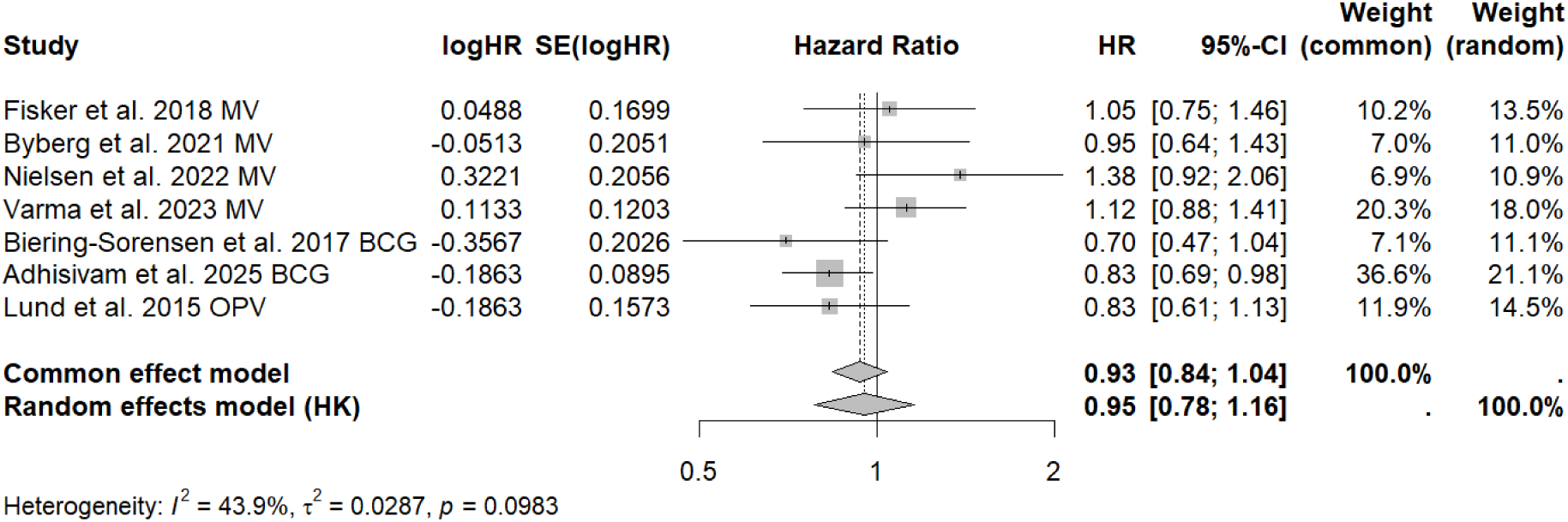
Forest plot of primary endpoint findings in 7 RCTs studying non-specific vaccine effects (NSEs) using all-cause mortality as the outcome, published during 2012-2025. Effect size (ES) is shown as point estimates and the 95% CIs are indicated with lines; ES under 1 indicates that the vaccine-intervention was beneficial; over 1 that it was harmful. The pooled effects analysis using the Hartung-Knapp-Sidik-Jonkman-method showed no significant effect of the interventions; random effects model showed an ES of 0.95, with a 95% CI 0.78-1.16 (corresponding to a vaccine effectiveness of 5% (range, −16% to 22%)).

The pooled analyses are shown as diamonds in the bottom of the plot. The common effects model yielded an ES of 0,93 (95% CI 0.84-1.04), while the random effects model, using the HKSJ-method, produced an ES of 0,95 (95% CI 0.78-1.16) (Figure 2).

As expected, the heterogeneity across the studies was moderate (I^2^=43.9%, t^2^=0.0287), indicating that approximately half the differences in the results are due to actual variations between studies (e.g., follow-up time, interventions, study designs), rather than sampling errors. This is apparent in the study-descriptions in Table 1 but can also be seen in figure 2 – the random effects model yields a slightly less precise estimate compared to the common effects model. Both 95% CIs included 1.0, indicating that across the included studies there was no statistically significant evidence to support the hypothesis of non-specific effects (NSEs) of vaccines on all-cause mortality.

## Discussion

### Summary of Findings – Evidence of NSE on mortality

We review the evidence for the non-specific effects (NSEs) of vaccines on all-cause mortality that has accumulated since a review commissioned by WHO-Sage from 2014 (4). To identify a causal NSE relationship, focusing on randomized controlled trials (RCTs) is essential. Since the 2014 WHO review, we identified seven additional RCTs that examined NSEs of live attenuated vaccines. In our meta-analysis, we pooled these, as the central hypothesis is always a dramatic reduction in all-cause mortality (1). Of the seven RCTs reviewed, only one set in India reported significant NSE, showing a 17% reduction (95% CI; 2-31%) in all-cause mortality among LBW children vaccinated with the BCG vaccine. Our meta-analysis of the primary endpoint across these seven trials found no significant evidence of NSEs on all-cause mortality. The overall effect size (ES) found a 5% reduction (95% CI; −16% to 22%) meaning a small non-significant NSE. We noted that the studies included in our analysis predominantly represents a single geographic setting (Guinea-Bissau), with only one study conducted outside this context, in India (21).

In October 2025, results from DTP trial conducted in 2012 in Guinea-Bissau were released, showing no significant NSEs of DTP on all-cause mortality (16% reduction, −37% to 48%) (30). This null finding was in preprint at the time our searches were completed and not included in our meta-analysis. Not including this paper did not affect our findings, because DTP is an inactivated vaccine and would not be included in our live-attenuated vaccine pool.

### Standardized Bias Assessment

Using the RoB-2 Cochrane Risk of Bias tool for RCTs, each of the 7 studies had “*some concern*” regarding bias especially due to the open-label design used. This is due to ethical constraints as the vaccines were already a part of the routine immunization schedule. These ethical constraints also prevented the use of double-blinding and placebo controls.

Several other methodological issues were identified: 1) promoting outcomes other than pre-specified primary endpoints; 2) adjustments for concurrent polio vaccination campaigns or reporting on a study period not listed as prespecified secondary outcomes in the clinical trial registry. Additionally, Støvring et al. (31) point out in their commentary that there is a high risk of randomly occurring false positives in analyses performed without corrections for multiple comparisons.

### Complete RCT evidence

Demonstrating NSEs in contemporary trials is particularly difficult because the vaccines in question are already included in the routine childhood EPI schedules. This limits the study designs to populations who have not yet received these vaccines as part of routine care, such as premature infants (where there is no recommendation to receive BCG at birth), or early measles doses before the standard immunization at 9-12 months in Africa. It also limits the use of placebo, out of concern that the mothers will think their children vaccinated (see for example (25,32)).

However, in the pre-EPI era (1940s to 1960s), several double-blinded placebo-controlled RCTs were conducted in Nigeria, Canada, and the United States to assess the effects of measles and BCG vaccines on the targeted diseases. In these trials, the cause of death was reviewed, allowing for the separation of target-disease-related deaths from deaths due to other causes (NSEs). Once we removed deaths attributable to the targeted disease, no statistically significant evidence of non-specific vaccine benefits was found. Notably, including the Nigerian study in their analysis, Higgins et al. had erroneously included measles-related deaths in the NSE effect estimate. We have since corrected this error in a secondary analysis and the accompanying plot (see figure 3).

**Figure 3.**
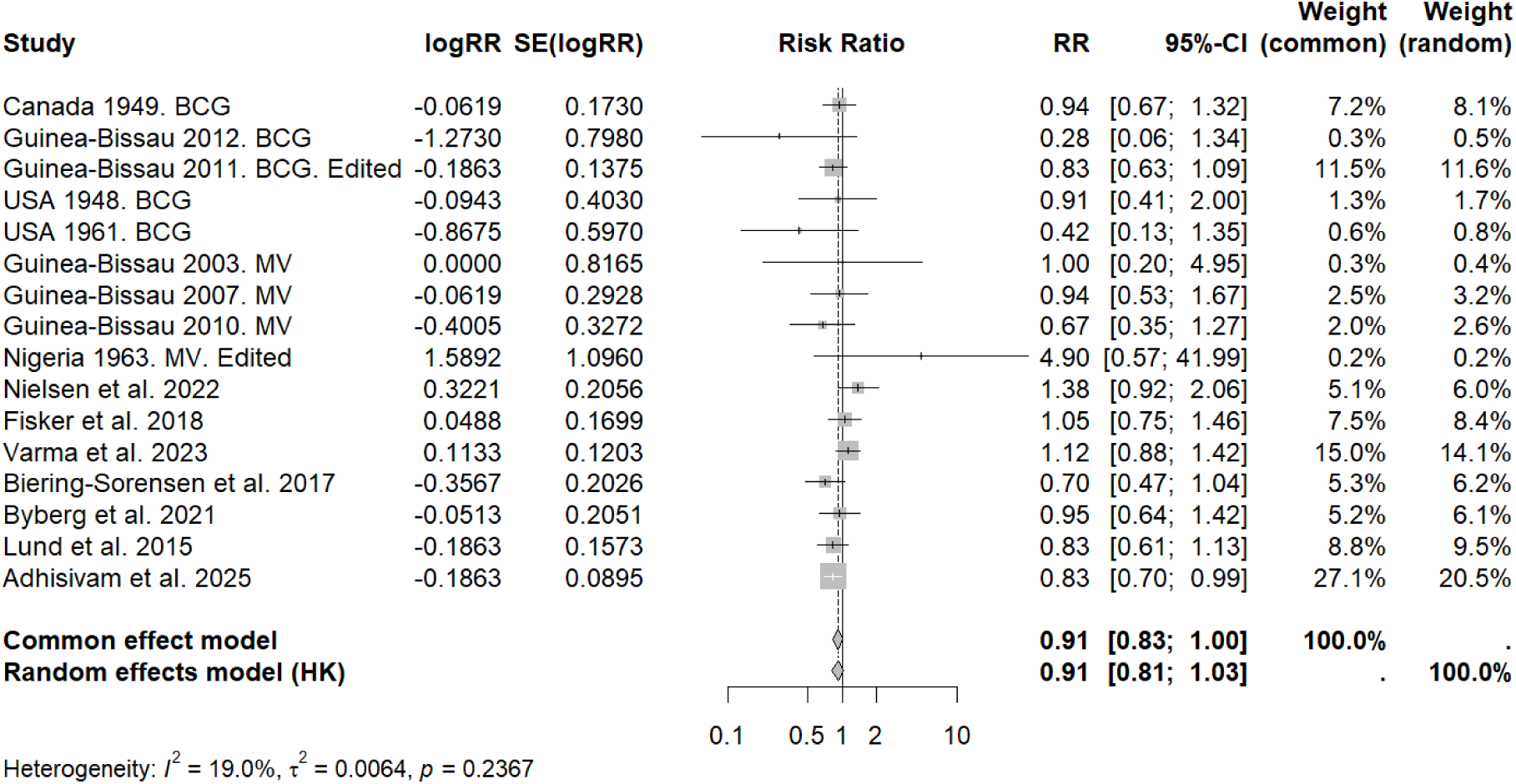
Edited data from the Nigerian study, as well as correcting the Guinea-Bissau 2011 study effect measure to the primary outcome – referenced as “edited” in the plot. The random effects model presents an ES of 0.91 with a 95% CI of 0.81-1.03. The overall effect trends in favor of the intervention; however, the effect is not statistically significant.

### A possible NSE mechanism

Interestingly, in the Indian RCT, the significant primary NSE finding appeared to be driven by a large difference in mortality in the first 3 days post-vaccination, a difference that was not found in the remainder of the 28-day follow-up period. A similar pattern was observed in several trials from Guinea-Bissau (1,21,33). This could suggest that NSEs (if any) are confined to the first 3 days after vaccination. Alternatively, this may signal issues with randomization in this open-label study design. The only way to resolve these uncertainties is through future placebo-controlled, double-blinded RCTs. In the Indian RCT, infection was synonymous with sepsis and *Klebsiella pneumoniae* was identified in 9.6% (7/73) of the isolates in the early vaccination group and 31% (41/133) in the control group; post hoc ratio 0.31 (95% CI 0.15; 0.66), p<0.001. However, *Klebsiella pneumoniae* is a common hospital acquired infection especially in intensive care units. Could it be, that the neonates receiving BCG and OPV received different care compared to the controls and the protection against sepsis in reality is a protection against hospital acquired infections? The study does not provide details of interventions such as intravenous catheters, oxygen supplementation (nasal tube or ventilator), nor the use of naso-gastric feeding, all of which could increase the risk of infection.

Evidence for NSE exists for measles where infections lead to immune amnesia due to T-cell depletion (34); such an effect would be expected to play out over months or several years. This effect on the adaptive immune system would take several weeks or months to appear and would last for years (35). If instead NSEs were due to innate immunity, the effects would happen immediately and be consistent with the observation of a reduction in the vaccine arm in the first days of the RCTs (9,21,22,33). We conducted a secondary analysis of NSEs stratified by the first 3 days versus the rest of the study period. In addition to the Indian study, Biering-Sørensen et al. (9) observed that six of sixteen deaths occurred within the first 3 days days after BCG vaccination. Also, we found that the ∼50% mortality reduction in the trial by Aaby et al. (22,32) was mostly occurring in the first 3 days (Table S4). These differences occur too early to be consistent with a T-cell depletion hypothesis.

### Caveats

One potential limitation of our analysis is that we pooled NSEs across all RCTs, regardless of vaccine, duration of the predefined study period, or type of effect measure (mortality rate ratio (MRR) vs. hazard ratio (HR)). This approach was based on the hypothesis that all live-attenuated vaccines was hypothesized to yield similar NSE reductions in all-cause mortality.

We adopted the treatment of MMR and HR as synonymous effect measures (effect size, ES) from Higgins et al. (4). Furthermore, the short primary follow-up time (28 days) in the only study utilizing MRR makes it a reasonable proxy for HR (22).

We also did not consider gender-specific NSEs i although this is also much discussed (36). We did this because gender-specific mortality was not a primary endpoint in the majority of the RCTs we reviewed, and the studies were therefore not randomized with respect to gender. Early observational studies suggested greater NSEs in girls, while more recent trials have shown stronger effects in boys or have failed to replicate any gender-based NSE differences (see for example 36,37).

Our review builds upon the WHO-commissioned review from 2014, which remains a cornerstone effort to evaluate the evidence base of NSEs of vaccines. The authors of the 2014 WHO review that examined all available evidence from both observational studies and RCTs, concluded that “*Randomized trials testing these hypotheses have been difficult or impossible to conduct on ethical grounds; as a result, with few exceptions, studies testing these hypotheses have been observational in nature*” (4). Furthermore, they noted that while RCTs on BCG vaccines tend to show a beneficial NSE, the findings are not conclusive. The review concluded that the available evidence does not provide conclusive support for NSE effects on all-cause mortality (4), and that the overall evidence base is insufficient to warrant changes in vaccine recommendations. The authors also called for additional RCT evidence, particularly regarding DTP vaccines.

For this reason, we revisited four historical trials conducted before the vaccines were in routine use (Table 2). These set out to study direct benefits of MV (measles) and BCG (tuberculosis) and are double-blinded and placebo-controlled studies (38–41). The pooled NSE effect across these 4 historical studies was in the random effects model (HKSJ-method) 0.99 (95% CI 0.41-2.42).

**Table 2.** Findings of Direct and Indirect (NSE) Vaccine Effects on Mortality in four “Historical” Placebo-Controlled and Blinded RCTs Conducted Before Measles and BCG Vaccines Were in Widespread Use. While designed to assess the direct vaccine effects, the cause-of-death data provided also allow for the calculation of indirect (NSE) effects. These four studies were also in the 2014 WHO-commissioned review (4)

| Country, year and intervention | N* | All deaths | Cause-specific deaths | Non-specific deaths | RR and VE Direct** | RR and VE indirect (NSE)** |
| --- | --- | --- | --- | --- | --- | --- |
| Nigeria, 1963*** (38), MV (measles) | 1962<br>I = 991<br>C = 971 | 17<br>I = 5<br>C = 12 | 11<br>I = 0<br>C = 11 | 6<br>I = 5<br>C = 1 | **0.04** (0.003-0.7)<br>**96%** (30-99,99%) | 4.90 (0.57-41.86)<br><0% (neg-43%) |
| Canada, 1949 (39), BCG (tuberculosis) | 606<br>I = 306<br>C = 303 | 116<br>I = 53<br>C = 63 | 11<br>I = 2<br>C = 9 | 105<br>I = 51<br>C = 54 | 0.22 (0.05-1.02)<br>78% (0-95%) | 0.94 (0.66-1.32)<br>6% (neg-34%) |
| USA, 1948 (40), BCG (tuberculosis) | 3006<br>I = 1551<br>C = 1457 | 164<br>I = 55<br>C = 109 | 59<br>I = 6<br>C = 53 | 105<br>I = 49<br>C = 56 | **0.11** (0.05-0.25)<br>**89%** (75-95%) | 0.83 (0.57-1.02)<br>17% (neg-43%) |
| USA, 1961**** (41), BCG (tuberculosis) | 451<br>I = 231<br>C = 220 | I = 4<br>C = 9 | I = 0<br>C = 4 | I = 4<br>C = 5 | 0.11 (0.006-1.95)<br>89% (neg-99%) | 0.76 (0.2-2.8)<br>23% (neg – 80%) |

\* I = intervention group, C = control group
\*\* Crude data available through articles. Relative risk is calculated using the MEDCALC calculator.
\*\*\* When calculating the NSE (indirect) relative risk for this study, we identified an error in the WHO-commissioned review: the estimate initially suggested an NSE (0.41; 95% CI 0.14-1.15) was based on a combined direct and indirect effect, including measles-related deaths. After excluding these deaths, the corrected estimate shows a non-significant negative NSE.
\*\*\*\* In this U.S.-based study among Indigenous populations in Southeast Alaska, the enrollment consisted of individuals under 20 years of age.

### Looking across all NSE evidence from RCTs

Our findings of no significant NSE across these trials contradict claims made by some authors who have promoted NSEs as well-established, paradigm-shifting phenomena of great importance to the EPI schedule (1). Instead, we find no evidence of a reduction in all-cause mortality across both the 7 trials published since 2012 and when adding 9 RCTs from the WHO review.

We have addressed the NSE question solely through the lens of all-cause mortality and found that claims of a major mortality reduction remain unsubstantiated. Few RCTs have demonstrated significant reductions in mortality attributable to vaccination beyond the expected protection against the targeted diseases. However, NSEs have been observed for less dramatic immunological or clinical effects, and these outcomes seem more appropriate going forward. The limitations of all-cause mortality as an endpoint are well documented in other fields. For example, a reported 50% reduction in all-cause mortality associated with influenza vaccination in elderly was later attributed to frailty bias (42–44). Given the inconsistent and methodologically fragile evidence for NSEs on all-cause mortality, it may be time to reconsider primary endpoints and refine the hypothesis to be tested.

### Going Forward

It is crucial to carefully evaluate the robustness of the evidence for NSEs on clinical endpoints. In secondary NSE literature (commentaries, reviews), the tendency to highlight positive findings from secondary endpoints, while little or no attention is given to primary negative findings is not helpful (see for example (36)). This concern of selective reporting has also been raised by other researchers (31).

Moreover, these authors cite findings from the WHO review as positive evidence of NSEs (REFs), when in fact Higgins et al. concluded that the NSE findings were only suggestive and that further RCT evidence was required to definitively address the issue (4). Such citations presenting secondary findings as NSE evidence causes confusion and does not reflect that the evidence for NSE reducing mortality remains controversial, at best.

As the controversy surrounding NSEs has now entered political discourse, possibly inspiring U.S. vaccine policy changes getting NSE right is more pertinent than ever. Settling the matter once and for all would require truly double-blinded, randomized placebo-controlled trials.

## Data Availability

All data used in the present study is derived from already published studies. Data and statistical programs is available in the manuscript.

## Acknowledgements

**Funding**

We acknowledge DNRF170 funding, the national research foundation of Denmark.

## References

1. Benn CS, Fisker AB, Rieckmann A, Sørup S, Aaby P. Vaccinology: time to change the paradigm? Lancet Infect Dis. 2020 Oct;20(10):e274–83.

2. Aaby P, Andersen M, Sodemann M, Jakobsen M, Gomes J, Fernandes M. Reduced childhood mortality after standard measles vaccination at 4-8 months compared with 9-11 months of age. BMJ. 1993 Nov 20;307(6915):1308–11.

3. Fine PEM, Williams TN, Aaby P, Källander K, Moulton LH, Flanagan KL, et al. Epidemiological studies of the ‘non-specific effects’ of vaccines: I – data collection in observational studies. Trop Med Int Health. 2009 Sept;14(9):969–76.

4. Higgins JPT, Soares-Weiser K, Reingold AL. Systematic review of the non-specific effects of BCG, DTP and measles containing vaccines [Internet]. WHO Strategic Advisory Group pf Experts on Immunization (SAGE); 2014. Available from: https://terrance.who.int/mediacentre/data/sage/SAGE_Docs_Ppt_Apr2014/9_session_non-specific_vaccine_effects/Apr2014_session9_epidemiologic_review.pdf

5. Higgins JPT, Soares-Weiser K, López-López JA, Kakourou A, Chaplin K, Christensen H, et al. Association of BCG, DTP, and measles containing vaccines with childhood mortality: systematic review. BMJ. 2016 Oct 13;i5170.

6. Page MJ, McKenzie JE, Bossuyt PM, Boutron I, Hoffmann TC, Mulrow CD, et al. The PRISMA 2020 statement: an updated guideline for reporting systematic reviews. Syst Rev. 2021 Dec;10(1):89.

7. Andersen A, Fisker AB, Rodrigues A, Martins C, Ravn H, Lund N, et al. National Immunization Campaigns with Oral Polio Vaccine Reduce All-Cause Mortality: A Natural Experiment within Seven Randomized Trials. Front Public Health. 2018;6:13.

8. Thysen SM, Fisker AB, Byberg S, Aaby P, Roy P, White R, et al. Disregarding the restrictive vial-opening policy for BCG vaccine in Guinea-Bissau: impact and cost-effectiveness for tuberculosis mortality and all-cause mortality in children aged 0-4 years. BMJ Glob Health. 2021 Aug;6(8).

9. Biering-Sørensen S, Aaby P, Napirna BM, Roth A, Ravn H, Rodrigues A, et al. Small Randomized Trial Among Low–birth-weight Children Receiving Bacillus Calmette-Guérin Vaccination at First Health Center Contact. Pediatr Infect Dis J. 2012 Mar;31(3):306–8.

10. Butkevičiūtė E, Jones CE, Smith SG. Heterologous effects of infant BCG vaccination: Potential mechanisms of immunity. Future Microbiol. 2018;13(10):1193–208.

11. Kjærgaard J, Birk NM, Nissen TN, Thøstesen LM, Pihl GT, Benn CS, et al. Nonspecific effect of BCG vaccination at birth on early childhood infections: a randomized, clinical multicenter trial. Pediatr Res. 2016 Nov;80(5):681–5.

12. Freyne B, Marchant A, Curtis N. BCG-associated heterologous immunity, a historical perspective: experimental models and immunological mechanisms. Trans R Soc Trop Med Hyg. 2015 Jan;109(1):46–51.

13. Freyne B, Marchant A, Curtis N. BCG-associated heterologous immunity, a historical perspective: intervention studies in animal models of infectious diseases. Trans R Soc Trop Med Hyg. 2015 Jan;109(1):52–61.

14. Benn CS, Roth A, Garly ML, Fisker AB, Schaltz-Buchholzer F, Timmermann CAG, et al. BCG scarring and improved child survival: a combined analysis of studies of BCG scarring. J Intern Med. 2020;288(6):614–24.

15. Benn CS, Fisker AB, Whittle HC, Aaby P. Revaccination with Live Attenuated Vaccines Confer Additional Beneficial Nonspecific Effects on Overall Survival: A Review. eBioMedicine. 2016 Aug;10:312–7.

16. Biering-Sørensen S, Jensen KJ, Monterio I, Ravn H, Aaby P, Benn CS. Rapid Protective Effects of Early BCG on Neonatal Mortality Among Low Birth Weight Boys: Observations From Randomized Trials. J Infect Dis. 2018 Feb 14;217(5):759–66.

17. Nielsen S, Fisker AB, Sié A, Müller O, Nebie E, Becher H, et al. Contradictory mortality results in early 2-dose measles vaccine trials: interactions with oral polio vaccine may explain differences. Int J Infect Dis [Internet]. 2024;148. Available from: https://www.scopus.com/inward/record.uri?eid=2-s2.0-85204392969&doi=10.1016%2Fj.ijid.2024.107224&partnerID=40&md5=ce09f81342aa570f4c005dbd4047697d

18. Trunk G, Davidović M, Bohlius J. Non-Specific Effects of Bacillus Calmette-Guérin: A Systematic Review and Meta-Analysis of Randomized Controlled Trials. Vaccines. 2023 Jan 4;11(1).

19. Aaby P, Martins CL, Ravn H, Rodrigues A, Whittle HC, Benn CS. Is early measles vaccination better than later measles vaccination? Trans R Soc Trop Med Hyg. 2015 Jan;109(1):16–28.

20. Aaby P, Nielsen S, Fisker AB, Pedersen LM, Welaga P, Hanifi SMA, et al. Stopping Oral Polio Vaccine (OPV) After Defeating Poliomyelitis in Low- and Middle-Income Countries: Harmful Unintended Consequences? Review of the Nonspecific Effects of OPV. Open Forum Infect Dis. 2022 Aug 2;9(8):ofac340.

21. Adhisivam B, Kamalarathnam C, Bhat BV, Jayaraman K, Namachivayam SP, Shann F, et al. Effect of BCG Danish and oral polio vaccine on neonatal mortality in newborn babies weighing less than 2000 g in India: multicentre open label randomised controlled trial (BLOW2). BMJ. 2025 Sept 22;390:e084745.

22. Biering-Sørensen S, Aaby P, Lund N, Monteiro I, Jensen KJ, Eriksen HB, et al. Early BCG-Denmark and Neonatal Mortality Among Infants Weighing <2500 g: A Randomized Controlled Trial. Clin Infect Dis. 2017 Oct 1;65(7):1183–90.

23. Lund N, Andersen A, Hansen ASK, Jepsen FS, Barbosa A, Biering-Sørensen S, et al. The Effect of Oral Polio Vaccine at Birth on Infant Mortality: A Randomized Trial. Clin Infect Dis. 2015 Nov 15;61(10):1504–11.

24. Fisker AB, Nebie E, Schoeps A, Martins C, Rodrigues A, Zakane A, et al. A Two-Center Randomized Trial of an Additional Early Dose of Measles Vaccine: Effects on Mortality and Measles Antibody Levels. Clin Infect Dis. 2018 May 2;66(10):1573–80.

25. Nielsen S, Fisker AB, Da Silva I, Byberg S, Biering-Sørensen S, Balé C, et al. Effect of early two-dose measles vaccination on childhood mortality and modification by maternal measles antibody in Guinea-Bissau, West Africa: A single-centre open-label randomised controlled trial. eClinicalMedicine. 2022 July;49:101467.

26. Byberg S, Aaby P, Rodrigues A, Stabell Benn C, Fisker AB. The mortality effects of disregarding the strategy to save doses of measles vaccine: a cluster-randomised trial in Guinea-Bissau. BMJ Glob Health. 2021 May;6(5):e004328.

27. Varma A, Thysen SM, Martins JSD, Nanque LM, Jensen AKG, Fisker AB. Overall effect of a campaign with measles vaccine on the composite outcome mortality or hospital admission: A cluster-randomized trial among children aged 9-59 months in rural Guinea-Bissau. Int J Infect Dis. 2023 Sept;134:23–30.

28. Benn CS, Fisker AB, Rodrigues A, Ravn H, Sartono E, Whittle H, et al. Sex-Differential Effect on Infant Mortality of Oral Polio Vaccine Administered with BCG at Birth in Guinea-Bissau. A Natural Experiment. Shea BJ, editor. PLoS ONE. 2008 Dec 29;3(12):e4056.

29. Higgins JPT, Savoric J, Page M, Elbers R, Sterne JAC. Assessing risk of bias in a randomized trial. In: Higgins J, Thomas J, Chandler J, Cumpston M, Li T, Page MJ, et al., editors. Cochrane handbook for systematic reviews of interventions. Second edition. Hoboken, NJ Chichester: Wiley Blackwell; 2019. p. 205–28. (Cochrane book series).

30. Agergaard J, Nielsen S, Benn CS, Aaby P. Randomised trial of not providing booster diphtheria-tetanus-pertussis (DTP) vaccination after measles vaccination and child survival: A failed trial [Internet]. Epidemiology; 2025 [cited 2025 Nov 27]. Available from: http://medrxiv.org/lookup/doi/10.1101/2025.10.29.25339081

31. Støvring H, Ekstrøm CT, Schneider JW, Strøm C. What is actually the emerging evidence about non-specific vaccine effects in randomized trials from the Bandim Health Project? Vaccine. 2025 Dec;68:127937.

32. Aaby P, Roth A, Ravn H, Napirna BM, Rodrigues A, Lisse IM, et al. Randomized Trial of BCG Vaccination at Birth to Low-Birth-Weight Children: Beneficial Nonspecific Effects in the Neonatal Period? J Infect Dis. 2011 July 15;204(2):245–52.

33. Moorlag SJCFM, Arts RJW, Van Crevel R, Netea MG. Non-specific effects of BCG vaccine on viral infections. Clin Microbiol Infect. 2019 Dec;25(12):1473–8.

34. Mina MJ. Measles, immune suppression and vaccination: direct and indirect nonspecific vaccine benefits. J Infect. 2017 June;74:S10–7.

35. Mina MJ, Metcalf CJE, De Swart RL, Osterhaus ADME, Grenfell BT. Long-term measles-induced immunomodulation increases overall childhood infectious disease mortality. Science. 2015 May 8;348(6235):694–9.

36. Aaby P, Netea MG, Benn CS. Beneficial non-specific effects of live vaccines against COVID-19 and other unrelated infections. Lancet Infect Dis. 2023 Jan;23(1):e34–42.

37. Sørensen MK, Schaltz-Buchholzer F, Jensen AM, Nielsen S, Monteiro I, Aaby P, et al. Retesting the hypothesis that early Diphtheria-Tetanus-Pertussis vaccination increases female mortality: An observational study within a randomised trial. Vaccine. 2022 Mar;40(11):1606–16.

38. Hartfield J, Morley D. Efficacy of measles vaccine. J Hyg (Lond). 1963 Mar;61(1):143–7.

39. Ferguson RG, Simes AB. BCG vaccination of Indian infants in Saskatchewan. Tubercle. 1949 Jan;30(1):5–11.

40. Aronson JD. Protective vaccination against tuberculosis with special reference to BCG vaccination. Am Rev Tuberc. 1948 Sept;58(3):255–81.

41. Rosenthal SR, Loewinsohn E, Graham ML, Liveright D, Thorne MG, Johnson V. BCG vaccination in tuberculous households. Am Rev Respir Dis. 1961 Nov;84:690–704.

42. Jackson LA, Jackson ML, Nelson JC, Neuzil KM, Weiss NS. Evidence of bias in estimates of influenza vaccine effectiveness in seniors. Int J Epidemiol. 2006 Apr 1;35(2):337–44.

43. Jackson LA, Nelson JC, Benson P, Neuzil KM, Reid RJ, Psaty BM, et al. Functional status is a confounder of the association of influenza vaccine and risk of all cause mortality in seniors. Int J Epidemiol. 2006 Apr 1;35(2):345–52.

44. Simonsen L, Viboud C, Taylor RJ, Miller MA. Mortality Benefits of Influenza Vaccination in the Elderly: An Ongoing Controversy. Am J Epidemiol. 2006 June 1;163(suppl_11):S168–S168.

