## Supplementary material. for "Non-specific effects of vaccines on all-cause mortality: a meta-analysis of randomized controlled trials (RCTs) 2012-2025"

#### Supplementary S1 – Sources and search strategy

##### Table S1.1 – Manual searches for RCTs on all-cause mortality:

| Search number | Date | Database | Search terms (all searches filtered for year 2012-2025) | Hits | Included for secondary screening | Reasons for exclusions | n (excl.) |
|---|---|---|---|---|---|---|---|
| 1 | 1/9/25 | SCOPUS | ("non specific effect*" OR "nonspecific effect*" OR "non-specific effect*" OR "heterologous effect*" OR "off-target effect*" OR "offtarget effect*" OR "off target effect*") AND (vaccin*) AND ("all cause mortality" OR "all-cause mortality" OR "allcause mortality") AND ("randomized controlled trial" OR rct) | 35 | Review Trunk_Bohlius 2023 | Analysis of more than one rct/metaanalysis/interrim analysis | 1 |
| | | | | | Review Aaby_Benn 2022 | Not RCT (observational, cohort, substudy, other methodology etc.) | 8 |
| | | | | | Nielsen_Benn 2022 | Not RCT (commentary, book chapter,, protocol, nonsystematic review, discussion etc.) | 8 |
| | | | | | Glynn_Fine 2021 | Not all-cause mortality as primary outcome (immological outcomes, respiratory illnesses, etc) | 4 |
| | | | | | Schaltz-Buchholzer_Benn 2020 | Not human study | 4 |
| | | | | | Fisker_Aaby 2018 | Not children < 5 | 1 |
| | | | | | Review Benn_Aaby 2016 | Trial discontinued | 1 |
| | | | | | Schaltz-Buchholzer_Benn 2024 | Not vaccination as intervention | 1 |
| 2 | 1/9/25 | SCOPUS | ("non specific effect*" OR "nonspecific effect*" OR "non-specific effect*" OR "heterologous effect*" OR "off-target effect*" OR "offtarget effect*" OR "off target effect*") AND (vaccin*) AND ("all cause mortality" OR "all-cause mortality" OR "allcause mortality") | 70 | Review Noble_Curtis 2023 | Not RCT (observational, cohort, substudy, other methodology etc.) | 15 |
| | | | | | (1)Review Trunk_Bohlius 2023 | Not RCT (commentary, book chapter,, protocol, nonsystematic review, discussion etc.) | 23 |
| | | | | | (1) Review Aaby_Benn 2022 | Analysis of more than one rct/metaanalysis | 1 |

| | | | | | | | |
|---|---|---|---|---|---|---|---|
| | | | | | (1) Nielsen_Benn 2022 | Not children < 5 | 1 |
| | | | | | (1)Glynn_Fine 2021 | Not human | 6 |
| | | | | | Review Benn_Aaby 2020 | Not all-cause mortality as primary outcome (immological outcomes, respiratory illnesses, etc) | 12 |
| | | | | | (1) Schaltz-Buchholzer_Benn 2020 | Trial discontinued | 1 |
| | | | | | (1) Fisker_Aaby 2018 | Not vaccination as intervention | 1 |
| | | | | | (1) Review Benn_Aaby 2016 | | |
| | | | | | Review Aaby_Benn 2015 | | |
| | | | | | (1) Schaltz-Buchholzer_Benn 2024 | | |
| 3 | 1/9/25 | Web of Science | ("non specific effect*" OR "nonspecific effect*" OR "non-specific effect*" OR "heterologous effect*" OR "off-target effect*" OR "offtarget effect*" OR "off target effect*") AND (vaccin*) AND ("all cause mortality" OR "all-cause mortality" OR “allcause mortality”) AND ("randomized controlled trial" OR rct) | 9 | (1) Schaltz-Buchholzer_Benn 2020 | Not RCT (observational, cohort, substudy, other methodology etc.) | 4 |
| | | | | | (1) Schaltz-Buchholzer_Benn 2024 | Not RCT (commentary, book chapter,, protocol, nonsystematic review etc.) | 1 |
| | | | | | | Not human | 2 |
| | | | | | | Not all-cause mortality as primary outcome (immological outcomes, respiratory illnesses, etc) | 1 |
| 4 | 1/9/25 | Web of Science | ("non specific effect*" OR "nonspecific effect*" OR "non-specific effect*" OR "heterologous effect*" OR "off-target effect*" OR "offtarget effect*" OR "off target effect*") AND (vaccin*) AND ("all cause mortality" OR "all-cause mortality" OR “allcause mortality”) | 47 | (1)Review Trunk_Bohlius 2023 | Not RCT (observational, cohort, substudy, other methodology etc.) | 14 |
| | | | | | (1) Nielsen_Benn 2022 | Not RCT (commentary, book chapter,, protocol, nonsystematic review etc.) | 11 |
| | | | | | (1) Review Benn_Aaby 2016 | Not human study | 6 |
| | | | | | (1)Glynn_Fine 2021 | Not all-cause mortality as primary outcome (immological outcomes, respiratory illnesses, etc) | 6 |

| | | | | | | | |
|---|---|---|---|---|---|---|---|
| | | | | | (1) Fisker_Aaby 2018 | Not children < 5 | 2 |
| | | | | | (2) Review Aaby_Benn 2015 | Not vaccination as intervention | 1 |
| | | | | | (1) Schaltz-Buchholzer_Benn 2020 | | |
| | | | | | (1) Schaltz-Buchholzer_Benn 2024 | | |
| 5 | 1/9/25 | EBSCOhost | ("non specific effect*" OR "nonspecific effect*" OR "non-specific effect*" OR "heterologous effect*" OR "off-target effect*" OR "offtarget effect*" OR "off target effect*") AND (vaccin*) AND ("all cause mortality" OR "all-cause mortality" OR "allcause mortality") AND ("randomized controlled trial" OR rct) | 15 | (1) Schaltz-Buchholzer_Benn 2020 | Not human | 2 |
| | | | | | (1) Fisker_Aaby 2018 | Not all-cause mortality as primary outcome | 2 |
| | | | | | (1)Review Trunk_Bohlius 2023 | Not RCT (observational, cohort, substudy etc.) | 4 |
| | | | | | (1) Review Benn_Aaby 2016 | Not RCT (commentary, book chapter,, protocol, nonsystematic review etc.) | 0 |
| | | | | | (1) Schaltz-Buchholzer_Benn 2024 | Not children < 5 | 2 |
| 6 | 1/9/25 | EBSCOhost | ("non specific effect*" OR "nonspecific effect*" OR "non-specific effect*" OR "heterologous effect*" OR "off-target effect*" OR "offtarget effect*" OR "off target effect*") AND (vaccin*) AND ("all cause mortality" OR "all-cause mortality" OR "allcause mortality") | 50 | (1)Glynn_Fine 2021 | Not human | 6 |
| | | | | | (1) Schaltz-Buchholzer_Benn 2020 | Not RCT (observational, cohort, substudy etc.) | 14 |
| | | | | | (1) Fisker_Aaby 2018 | Not RCT (commentary, book chapter,, protocol, nonsystematic review etc.) | 11 |
| | | | | | (1)Review Trunk_Bohlius 2023 | Analysis of more than one rct/metaanalysis | 1 |
| | | | | | (1) Nielsen_Benn 2022 | Not all-cause mortality as primary outcome | 7 |
| | | | | | (1) Review Benn_Aaby 2016 | Not children < 5 | 2 |
| | | | | | (2) Review Aaby_Benn 2015 | Not vaccination as intervention | 1 |
| | | | | | (1) Schaltz-Buchholzer_Benn 2024 | Not human | 2 |

| | | | | | | | |
|---|---|---|---|---|---|---|---|
| | | | | | | Not RCT (observational, cohort, substudy, other methodology etc.) | 2 |
| 7 | 2/9/25 | PubMed | ("non specific effect*" OR "nonspecific effect*" OR "non-specific effect*" OR "heterologous effect*" OR "off-target effect*" OR "offtarget effect*" OR "off target effect*") AND (vaccin*) AND ("all cause mortality" OR "all-cause mortality" OR “allcause mortality”) AND ("randomized controlled trial" OR rct) | 13 | (1)Glynn_Fine 2021 | Not RCT (commentary, book chapter,, protocol, nonsystematic review etc.) | 1 |
| | | | | | (1) Schaltz-Buchholzer_Benn 2020 | Not human | 2 |
| | | | | | (1) Fisker_Aaby 2018 | Not all-cause mortality as primary outcome | 1 |
| | | | | | (1) Schaltz-Buchholzer_Benn 2024 | Not RCT (observational, cohort, substudy, other methodology etc.) | 2 |
| 8 | 2/9/25 | PubMed | ("non specific effect*" OR "nonspecific effect*" OR "non-specific effect*" OR "heterologous effect*" OR "off-target effect*" OR "offtarget effect*" OR "off target effect*") AND (vaccin*) AND ("all cause mortality" OR "all-cause mortality" OR "allcause mortality") | 49 | (1)Review Trunk_Bohlius 2023 | Not RCT (commentary, book chapter,, protocol, nonsystematic review etc.) | 11 |
| | | | | | (2) Review Aaby_Benn 2015 | Not all-cause mortality as primary outcome | 6 |
| | | | | | (1)Glynn_Fine 2021 | Not human | 6 |
| | | | | | (1) Nielsen_Benn 2022 | Not RCT (observational, cohort, substudy, other methodology etc.) | 16 |
| | | | | | (1) Review Benn_Aaby 2016 | Not children < 5 | 1 |
| | | | | | (1) Schaltz-Buchholzer_Benn 2020 | Not vaccination as intervention | 1 |
| | | | | | (1) Fisker_Aaby 2018 | | |
| | | | | | (1) Schaltz-Buchholzer_Benn 2024 | | |
| 9 | 2/9/25 | Cochrane | ("non specific effect*" OR "nonspecific effect*" OR "non-specific effect*" OR "heterologous effect*" OR "off-target effect*" OR "offtarget effect*" OR "off target effect*") AND (vaccin*) AND ("all cause mortality" OR "all-cause mortality" OR "allcause mortality") | 14 trials | (1)Glynn_Fine 2021 | Not RCT (observational, cohort, substudy, other methodology etc.) | 4 |

| | | | | | | | |
|---|---|---|---|---|---|---|---|
| | | | | | Varma_Fisker 2023 | Not all-cause mortality as primary outcome | 4 |
| | | | | | (1) Nielsen_Benn 2022 | | |
| | | | | | (1) Review Benn_Aaby 2016 | | |
| | | | | | (1) Fisker_Aaby 2018 | | |
| | | | | | (1) Schaltz-Buchholzer_Benn 2024 | | |
| 10 | 2/9/25 | Cochrane | ("non specific effect*" OR "nonspecific effect*" OR "non-specific effect*" OR "heterologous effect*" OR "off-target effect*" OR "offtarget effect*" OR "off target effect*") AND (vaccin*) AND ("all cause mortality" OR "all-cause mortality" OR "allcause mortality") AND ("randomized controlled trial" OR rct) | 14 | (1)Glynn_Fine 2021 | Not RCT (observational, cohort, substudy, other methodology etc.) | 5 |
| | | | | | (11) Varma_Fisker 2023 | Not all-cause mortality as primary outcome | 4 |
| | | | | | (1) Nielsen_Benn 2022 | | |
| | | | | | (1) Review Benn_Aaby 2016 | | |
| | | | | | (1) Schaltz-Buchholzer_Benn 2024 | | |
| 11 | 2/9/25 | Trunk:_Bohlius review | Chainsearch | 20 | Biering-Sørensen_Benn 2017 | Published before 2012 | 2 |
| | | | | | Kjærgaard_Stensballe 2016 | Not all-cause mortality as primary outcome | 7 |
| | | | | | Jayaraman_Vishnu 2019 | Included in WHO review | 1 |
| | | | | | (1)Glynn_Fine 2021 | Not children < 5 | 6 |
| 12 | 2/9/25 | Aaby_Benn 2022 | Chainsearch | 4 rcts | Byberg_Fisker 2021 | Analysis of more than one rct/metaanalysis/interrim analysis | 1 |
| | | | | | Varma_Fisker 2022 | | |
| | | | | | Nielsen_ 2022 | | |

| | | | | | | | |
|---|---|---|---|---|---|---|---|
| 13 | 2/9/25 | Benn_Aaby 2016 | Chainsearch | 2 rcts | | Published before 2012 | 2 |
| 14 | 3/9/25 | Aaby_Benn 2015 | Chainsearch | 4 rcts | | Published before 2012 | 4 |
| 15 | 3/9/25 | Noble_Curtis 2023 | Chainsearch | 11 rcts | | Not children < 5 | 11 |
| 16 | 3/9/25 | Benn_Aaby 2020 | Chainsearch | 5 RCTs | Berendsen_Aaby 2020 | Published before 2012 | 1 |
| | | | | | Lund_ 2015 | Not RCT (observational, cohort, substudy, other methodology etc.) | 1 |
| | | | | | | Not vaccination as intervention | 1 |
| 17 | 13/10/25 | SCOPUS | ("non specific effect*" OR "nonspecific effect*" OR "non-specific effect*" OR "heterologous effect*" OR "off-target effect*" OR "offtarget effect*" OR "off target effect*") AND (vaccin*) AND ("all cause mortality" OR "all-cause mortality" OR “allcause mortality”) AND ("randomized controlled trial" OR rct) | 36 | Adhisivam_Sundaram 2025 | Analysis of more than one rct/metaanalysis/interrim analysis | 1 |
| | | | | | (1) Review Trunk_Bohlius | Not RCT (observational, cohort, substudy, other methodology etc.) | 8 |
| | | | | | (1) Review Aaby_Benn 2022 | Not children < 5 | 2 |
| | | | | | (1) Fisker_Aaby 2018 | Not all-cause mortality as primary outcome | 6 |
| | | | | | (1) Review Benn_Aaby 2016 | Not RCT (commentary, book chapter,, protocol, nonsystematic review etc.) | 8 |
| | | | | | | Not human | 4 |
| | | | | | | Trial discontinued | 1 |

### Table S1.2 – Full text screening of articles

This table presents the manual full text screening. Red background marks reason for exclusion. Green marks included in this review.

| Author | Year | Title | Trial registration | Primary outcome from clinical trial registration | Primary outcome according to article | Secondary outcome according to article | Intervention | Control | Time of intervention | Age group | Effect estimate |
|---|---|---|---|---|---|---|---|---|---|---|---|
| Schaltz-Buchholzer et al. | 2024 | Effects of Neonatal BCG-Japan Versus BCG-Russia Vaccination on Overall Mortality and Morbidity: Randomized Controlled Trial From Guinea-Bissau (BCGSTRAIN II) | NCT03400878 | Hospital admissions at 6 weeks | All-cause hospital admissions | | | | | | |
| Trunk, Davidovic & Bohluis | 2022 | Non-specific effects of bacillus calmette-Guérin: a systematic review and meta-analysis of randomized controlled trials | CRD 42021255017 | REVIEW (see search protocol) | | | | | | | |
| Aaby et al. | 2022 | Stopping oral polio vaccine (OPV) after defeating poliomyelitis in low-and middle-income coundtries: harmful unintended consequenses.Review of the nse of opv | n.a. | REVIEW | | | | | | | |
| Benn et al. | 2016 | Revaccination with live attenuated vaccines confer additional beneficial nonspecific effects on overall survival: a review | n.a. | REVIEW | | | | | | | |
| Benn et al. | 2020 | BCG scarring and improved child survival: a combined analysis of studies of bcg scarring. | n.a. | REVIEW | | | | | | | |
| Glynn et al. | 2021 | The effect of BCG revaccination on all-cause mortality beyond infancy: 30-years follow-up of a population-based, double-blind, randomized placebo-controlled trial in Malawi | n.a. | Outcomes of leprosy and tuberculosis | All-cause mortality | | | | | 3 months-75 years | |
| Schaltz-Buchholzer et al. | 2020 | Early Vaccination with Bacille Calmette-Guérin-Denmartk or BCG-Japan Versus BCG-Russia to Healthy | NCT02447536 | Hospital admissions at 6 weeks | all-cause hospital admission within 6 weeks after birth | Neonatal admissions, mortality by 6 weeks, bcg skin reaction frequency, etc. | BCG Denmark | BCG-Russia | At birth before hospital discharge | | |

| | | | | | | | | | | | |
|---|---|---|---|---|---|---|---|---|---|---|---|
| | | Newborns in Guinea-Bissau: A Randomized Controlled Trial | | | | | | | | | |
| Kjærgaard et al. | 2016 | Nonspecific effect of BCG vaccination at birth on early vhildhood infections: a randmized, clinical multicenter trial | NCT01694108 | All-cause hospitalization 0-15 months | No of infectious illness episodes from 0-3 mo and 3-13 mo | No of visits to the GP | | | | | |
| Jayaraman et al. | 2019 | Two randomized trials of the effects of the Russian strain of BCG alone or with polio vaccine on neonatal mortality in infants weighing <2000g in India | n.a. | | In-hospital death up to 28 days | Overall mortality up to 28 days | BCG-Russia & BCG with OPV | Following normal program - normally after child gains weight/is discharged from NICU | As soon as possible after admission to NICU and within 48 hours | | |
| Varma et al. | 2022 | Research protocol of two concurrent cluster-randomized trials: Real-life effect of CAMPaign with measles vaccination on mortality and morbidity amon children in rural Guinea-Bissau | NCT03460002 | | | | | | | | |
| Nielsen et al. | 2022 | Effect of early two-dose measles vaccination on childhood mortality and modification by maternal measles antibody in Guinea-Bissau, West Africa: A single-centre open-label randomised controlled trial | DUPLICATE. Nielsen et al. 2022 is included. | | | | | | | | |
| Berendsen et al. | 2020 | Maternal Priming: Bacillus Calmette-Guérin (BCG) Vaccine Scarring in Mothers Enhances the Survival of Their Child With a BCG Scar | | Mortality | | | | | | | |
| Nielsen et al. | 2022 | Effect of early two-dose measles vaccination on childhood mortality and modification by maternal measles antibody in Guinea-Bissau, West Africa: A single-centre open-label randomized controlled trial | NCT01486355. | Mortality | All-cause mortality | | MV at 4 months + 9 months | MV at 9 months | at 4 months | Children 4-7.5 months of age | HR |
| Fisker et al. | 2018 | A Two-Center Randomized Trial of an Additional Early dose of Measles vaccine: Effects on mortality and mealses antibody levels | NCT01644721 | Mortality 4m-3y | All-cause mortality | levels of measles antibody | Early MV | No early MV | 4 weeks after third Penta vaccine | 121-215 days | HR |
| Varma et al. | 2023 | Overall effect of a campaign with measles vaccine on the composite outcome mortality or hospital | NCT03460002 | Composite outcome: mortality and hospital admission (follow up, up to two years) | Composite outcome: mortality and non-accidental | mortality, hospital admission, cause-specific mortality/hospital admission | MV (or OPV according to age of enrollment) and a health check-up | Health check-up | vaccination at enrollment | Children 9-59 months (Healthy) | HR |

| | | | | | | | | | | | |
|---|---|---|---|---|---|---|---|---|---|---|---|
| | | admission: a cluster-randomized trial among children aged 9-59 months in rural Guinea-Bissau | | | hospital admission | | | | | | |
| Biering-Sørensen et al. | 2017 | Early BCG-Denmark and neonatal mortality among infants weighing <2500 g: A randomized controlled trial | NCT00625482 | Mortality | Neonatal mortality within 28 days of life | infant mortality, within 12 months of life | BCG early | BCG later (mothers were encouraged to have infant vaccinated, when they had gained weight) | At birth | Neonates weighing <2500 g | MRR |
| Byberg et al. | 2021 | The mortality effects of disregarding the strategy to save doses of measles vaccine: a cluster randomized trial in Guinea-Bissau | NCT01306006 | Mortality | Mortality | | MV for all (children aged 9-35 month | Restrictive MV-policy (children aged 9-11, if 6 or more were present at the same time | At enrollment | 9-35months | HR |
| Lund et al. | 2015 | The effect of oral polio vaccine at birth on mortality. A randomized clinical trial | NCT00710983 | Mortality by sex | Overall mortality within the first 12 months | | OPV + BCG at randomization | BCG only | At randomization/enrollment | Neonates, healthy | HR |
| Adhisivam et al. | 2025 | Effect of BCG Danish and oral polio vaccine on neonatal mortality in newborn babies weighing less than 2000 g in India: multicentre open label randomised controlled trial (BLOW2) | CTRI/2017/01/007676 | Neonatal mortality | All-cause neonatal mortality | Neonatal mortality due to infection | BCG (+ OPV) at enrollment | BCG (+ OPV later) | At enrollment | LBW Neonates weighing <2000 g | HR |

#### Supplementary S2 – Statistical analysis

**Common and random effect model, meta-analysis used in R meta.**

We used the following code in R utilizing the meta package. We used Hartung-Knapp-Sidik-Jonkman (HKSJ) method to estimate confidence intervals, to account for the small number of studies as well as between-study heterogeneity.

```
library(meta)

study_a = c("Fisker et al. 2018 MV",
            "Byberg et al. 2021 MV",
            "Nielsen et al. 2022 MV",
            "Varma et al. 2023 MV",
            "Biering-Sorensen et al. 2017 BCG",
            "Adhisivam et al. 2025 BCG",
            "Lund et al. 2015 OPV")
ES_a    = c(1.05, 0.95, 1.38, 1.12, 0.70, 0.83, 0.83)
lower_a = c(0.75, 0.64, 0.92, 0.88, 0.47, 0.69, 0.61)
upper_a = c(1.46, 1.43, 2.06, 1.41, 1.04, 0.98, 1.13)
logHR_a <- log(ES_a)
selogHR_a <- (log(upper_a) - log(lower_a)) / (2 * 1.96)
meta_hr_a <- metagen(TE = logHR_a, seTE = selogHR_a, studlab = study_a, sm = "HR", method.tau = "SJ", hakn=TRUE,
hakn.adjust=TRUE)
meta_hr_a$lower <- log(lower_a)
meta_hr_a$upper <- log(upper_a)
summary(meta_hr_a)
forest_mort <- forest(meta_hr_a)
```

### Supplementary S3 – standardized bias assessment using the Cochrane Risk of Bias 2 (RoB-2) tool

Risk of bias was assessed using the Cochrane RoB 2 tool, which evaluates potential bias in randomized controlled trials (RCTs) across five domains. In Figure S3, the assessments we made of the seven included studies are presented. Three studies were per-protocol analyses, whereas four studies were intention-to-treat analyses, as shown in the figure.

**Per-protocol**

| Unique ID | Study ID | Experimental | Comparator | Outcome | Weight | D1 | D2 | D3 | D4 | D5 | Overall |
|---|---|---|---|---|---|---|---|---|---|---|---|
| Biering-Sørens | Biering-Sørensen e | BCG early | BCG later | All-cause mortality (MF | 1 | ! | ! | + | + | + | ! |
| Nielsen et al., 2 | Nielsen et al., 2022 | MV at 4 months | MV at 9 months | All-cause mortality (HR | 1 | ! | ! | + | + | + | ! |
| Fisker et al., 2 | Fisker et al., 2018 | Early MV | No early MV | All-cause mortality (HR | 1 | + | ! | + | + | + | ! |

| | |
|---|---|
| + | Low risk |
| ! | Some concerns |
| - | High risk |

| | |
|---|---|
| D1 | Randomisation process |
| D2 | Deviations from the intended interventions |
| D3 | Missing outcome data |
| D4 | Measurement of the outcome |
| D5 | Selection of the reported result |

**Intention-to-treat**

| Unique ID | Study ID | Experimental | Comparator | Outcome | Weight | D1 | D2 | D3 | D4 | D5 | Overall |
|---|---|---|---|---|---|---|---|---|---|---|---|
| Varma et al., 2 | Varma et al., 2023 | MV + health check up | Health check up | All-cause mortality me | 1 | ! | ! | + | + | + | ! |
| Lund et al., 20 | Lund et al., 2015 b | OPV+BCG at randomizati | BCG only | All-cause mortality (HR | 1 | ! | ! | + | + | + | ! |
| Byberg et a., 2 | Byberg et al., 2021 | MV for all | Restrictive MV | All-cause mortality (HR | 1 | ! | ! | + | + | + | ! |
| Adhisivam et a | Adhisivam et al., 2 | BCG+OPV early | BCG + OPV later | All-cause mortality (HR | 1 | + | ! | + | + | + | ! |

| | |
|---|---|
| + | Low risk |
| ! | Some concerns |
| - | High risk |

| | |
|---|---|
| D1 | Randomisation process |
| D2 | Deviations from the intended interventions |
| D3 | Missing outcome data |
| D4 | Measurement of the outcome |
| D5 | Selection of the reported result |

**Figure S3** – RoB 2 assessments of the seven included articles.

#### Supplementary 4 – Additional analyses

**Timing of the effect: first three days versus the rest of the study period.**

Three of the seven RCTs we included in this study, and a 2011 and 2012 study allowed stratification of the first three days and the rest of the study period. All studies were conducted with neonates (healthy, or low birthweight (LBW)). This table shows the relative risk measures of all-cause mortality over time. The relative risk estimates were either directly available in the publications, or else computed from tables and figures by us. As this is a secondary finding, it should be interpreted as such.

| **Study, Follow up, and Effect Measure** | **Entire Study Period (pre-specified primary endpoint)** | First 3 days of the study period (immediately after vaccination) | Remaining study period (after day 3) |
|---|---|---|---|
| **Adhisivam et al 2025** (21)<br>**28 days**<br>**HR** (95% CI) | **0.83**<br>(0.69-0.98) | 0.72<br>(0.51-1.03) | 0.92<br>(0,76-1,12) |
| **Biering-Sørensen et al 2017** (22)<br>**Neonatal period (28 days)**<br>**MMR (**95% CI) | 0.70<br>(0.47-1.04) | 0.68<br>(0.33-1.40) | **0.77**<br>(0.60-0.98) |
| **Lund et al 2015** (23)<br>**12mo**<br>**HR** (95% CI) | 0.83<br>(0.61-1.13) | **0,58**<br>(0,38-0,90) | 1,26<br>(0,79-2,00) |
| **Biering-Sørensen et al 2012** (9)<br>**12 mo**<br>**MMR** (95% CI) | 0.41<br>(0.14-1.18) | 0.17<br>(0.02-1.35) | 0.86<br>(0.25-3.04) |
| **Aaby et al 2011** (32)<br>**12 mo**<br>**MMR** (95% CI) | 0.83<br>(0.63-1.08) | 0.49<br>(0.21-1.15) | 0.89<br>(0.68-1.15) |

**Table S4** – Timing of the non-specific effects of vaccines

#### Pooled effect of from all live-attenuated vaccines

This figure presents the pooled effect from both data identified through our review as well as uncorrected data as it appears in the WHO-commissioned review. This crude data was conducted as a meta-analysis in *R* using the *meta* package, with random effects model fitted using Hartung-Knapp adjustment and restricted maximum likelihood (REML).

The corrections done in Figure 3 does not affect the pooled effect by much – ES 0.88 (95% CI 0.76-1.02) in Figure S4, and ES 0.91 (95% CI 0.81-1.03) in Figure 3. The corrections included in Figure 3 was correcting for the primary outcome (Guinea-Bissau 2011) and effect size (Nigeria 1963).

| Study | logRR | SE(logRR) | RR | 95%-CI | Weight (common) | Weight (random) |
|---|---|---|---|---|---|---|
| Canada 1949. BCG | -0.0619 | 0.1730 | 0.94 | [0.67; 1.32] | 7.8% | 9.1% |
| Guinea-Bissau 2012. BCG | -1.2730 | 0.7980 | 0.28 | [0.06; 1.37] | 0.4% | 0.6% |
| Guinea-Bissau 2011. BCG | -0.5978 | 0.2455 | 0.55 | [0.34; 0.89] | 3.9% | 5.5% |
| USA 1948. BCG | -0.0943 | 0.4030 | 0.91 | [0.41; 1.99] | 1.4% | 2.4% |
| USA 1961. BCG | -0.8675 | 0.5970 | 0.42 | [0.13; 1.35] | 0.7% | 1.1% |
| Guinea-Bissau 2003. MV | 0.0000 | 0.8165 | 1.00 | [0.20; 4.91] | 0.4% | 0.6% |
| Guinea-Bissau 2007. MV | -0.0619 | 0.2928 | 0.94 | [0.53; 1.67] | 2.7% | 4.1% |
| Guinea-Bissau 2010. MV | -0.4005 | 0.3272 | 0.67 | [0.33; 1.19] | 2.2% | 3.4% |
| Nigeria 1963. MV | -0.8916 | 0.5438 | 0.41 | [0.14; 1.18] | 0.8% | 1.4% |
| Nielsen et al. 2022 | 0.3221 | 0.2056 | 1.38 | [0.92; 2.06] | 5.5% | 7.2% |
| Fisker et al. 2018 | 0.0488 | 0.1699 | 1.05 | [0.75; 1.46] | 8.1% | 9.3% |
| Varma et al. 2023 | 0.1133 | 0.1203 | 1.12 | [0.88; 1.41] | 16.2% | 13.5% |
| Biering-Sorensen et al. 2017 | -0.3567 | 0.2026 | 0.70 | [0.47; 1.04] | 5.7% | 7.3% |
| Byberg et al. 2021 | -0.0513 | 0.2051 | 0.95 | [0.64; 1.43] | 5.6% | 7.2% |
| Lund et al. 2015 | -0.1863 | 0.1573 | 0.83 | [0.61; 1.13] | 9.5% | 10.2% |
| Adhisivam et al. 2025 | -0.1863 | 0.0895 | 0.83 | [0.69; 0.98] | 29.2% | 17.0% |
| **Common effect model** | | | **0.89** | **[0.81; 0.98]** | **100.0%** | . |
| **Random effects model (HK)** | | | **0.88** | **[0.76; 1.02]** | . | **100.0%** |

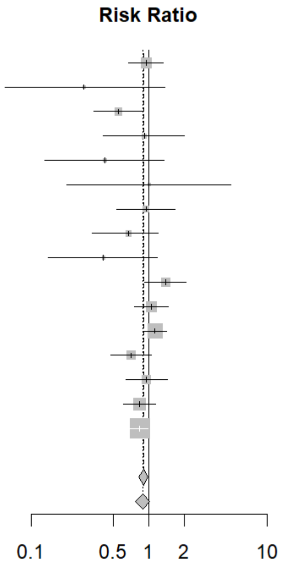

Heterogeneity: $I^2 = 31.6\%$, $\tau^2 = 0.0169$, $p = 0.1099$

**Figure S4** – meta-analysis of 16 studies on non-specific effects of vaccines on all-cause mortality.
